# Automated Chest Radiographs Triage Reading by a Deep Learning Referee Network

**DOI:** 10.1101/2021.06.01.21257399

**Authors:** Rafael López-González, Jose Sánchez-García, Belén Fos-Guarinos, Fabio García-Castro, Ángel Alberich-Bayarri, Emilio Soria-Olivas, Carlos Muñoz-Núñez, Luis Martí-Bonmatí

**Affiliations:** Quantitative Imaging Biomarkers in Medicine (Quibim S.L.). Valencia, Spain; Intelligent Data Analysis Laboratory (IDAL). ETSE. Universidad de Valencia. Valencia, Spain; Universidad Politécnica de Valencia. Valencia, Spain; Department of Radiology and GIBI230 Research Group on Biomedical Imaging. Hospital Universitario y Politécnico de La Fe and Instituto de Investigación Sanitaria La Fe. Valencia, Spain

**Keywords:** Deep Learning, Machine Learning, Chest Radiographs, Computer-Aided Diagnosis, Convolutional Neural Networks

## Abstract

Chest radiographs are often obtained as a screening for early diagnosis tool to rule out abnormalities mainly related to different cardiovascular and respiratory diseases. Reading and reporting numerous chest radiographs is a complex and time-consuming task. This research proposes and evaluates a deep learning (DL) approach based on convolutional neural networks (CNN) combined with a referee fully connected neural network as a computer-aided diagnosis tool in chest X-ray triage and worklist prioritization. The CNN models were trained with a combination of three large scale databases: ChestX-ray14, CheXpert and PadChest. The final database contained 327,176 images labeled with findings obtained by natural language processing (NLP) techniques applied to the radiology reports. The dataset was split in 16 different balanced binary partitions, which were used to train 16 finding-specific classification CNNs. Afterwards, a normal vs abnormal partition of the dataset was created, being abnormal the presence of at least one pathologic change. This final partition was used to train a fully connected neural network as referee that was fed with all the 16 previously trained outcomes. The Area Under the Curve (AUC) analysis evaluated and compared the performance of the models. The system was successfully implemented and evaluated with a test set of 3400 images. The AUC of the normal vs abnormal classification was 0.94. The highest AUC of the finding-specific classifiers was 0.99 for hernia. The proposed system can be used to assist radiologists identifying abnormal exams, allowing a time-efficiency triage approach.

## 1. Introduction

Chest plain films are used worldwide in the evaluation of patients with a clinically variable cardio-respiratory disease expectancy. Chest radiographs represent one of the largest workloads in radiology departments. Cardiovascular and respiratory diseases are considered a global health problem, a challenge for our health care systems, having a huge socio-economic impact. An early evaluation of these disorders can be considered imperative (World Health Organization, 2017) (World Health Organization, 2011) (European Respiratory Society, 2013).

Despite the technological advancement in CT imaging, chest plain radiographs remain the most frequently performed diagnostic examination technique in daily practice (Bayo et al., 2005) including emergency care. Chest radiographs provide information on several relevant and sometimes life-threatening cardiovascular and respiratory diseases, providing data on anatomical structures such as the heart, lungs and bones. Radiographs are a cost-effective procedure when compared to other imaging techniques.

Chest radiographs are complex to read and time consuming, requiring a large learning curve (Eisen et al., 2006) (Herman & Hessel, 1975). Unfortunately, radiologists are scarce for this workflow. Increase number of radiological reports per working day, and lack of professionals in large heavily loaded centers and in rural areas or economically underdeveloped countries, are some of the reasons related to this manpower shortage (Mollura et al., 2010). Absence of enough radiologists and overflow of heavy daily work led to potential errors, misdiagnosis and burnouts, directly related with an increased morbidity and mortality (Kesselman et al., 2016). Therefore, it seems evident that an automatic triage system for disease detection in chest X-ray films would have an important value for patients, clinicians and consequently, health care organizations. This system would help radiologists to prioritize either reading exams with most probable abnormalities or avoiding reports when the probability of having an abnormality is extremely low.

Computer-aided diagnosis (CAD) systems have experienced a dramatic progress over the last decade, mostly since 2015 (van Ginneken, 2017). Currently, we are witnessing a revolution of Deep Learning (DL) solutions on medical imaging due to the exponential growth of the amounts of curated data available on the net and the wide availability of graphical processing units (GPUs), making parallel processing faster and cheaper (Hwang, 2018).

Nowadays, the main challenge to develop a DL CAD system for chest radiographs triage is data scarcity. To develop DL solutions, a very large dataset that contains examples of all possible situations, including different pathologies and whole disease spectra that can be found in a chest radiograph, is necessary. To help researchers, the NIH released ChestX-ray14, a chest X-ray multi-label dataset which encloses 112,120 posterior-anterior chest exams labeled with 14 different radiological findings (Wang et al., 2017). This database boosted the research of DL applications for chest X-ray analysis using end-to-end CNNs for multi-label classification (Rajpurkar, 2017). Following the same path, CheXpert (Irvin et al., 2017) and PadChest (Bustos et al., 2020) databases were made public with 224,316 radiographs labeled with 14 findings and 160,000 radiographs labeled with 174 findings respectively. Due to the unequal occurrence of the different pathologies in patients, there is a large class imbalance in the datasets, which was not considered in previous approaches and could affect negatively the performance of CNNs (Buda et al., 2017). Therefore, a training methodology designed to face this problem is needed.

Our objective is to present a new DL solution to assist radiologists by triaging chest radiographs, providing an output of either normal or abnormal, abnormal meaning that cardiovascular, respiratory diseases or fractures have been detected. A novel classification algorithm was trained with ChestX-ray14, CheXpert and PadChest balanced partitions using a combination of pathology-specific CNNs to provide the likelihood of a plain chest exam to be abnormal by means of a referee fully connected neural network.

## 2. Materials and methods

### 2.1. Database prepation

The large-scale public databases ChestX-ray14 (Wang et al., 2017), CheXpert (Rajpurkar, 2017) and PadChest (Bustos et al., 2020) were combined to maximize the data for training the model. The resulting database included 327,176 radiographs of which 94,334 proceeded from ChestX-ray14, 192,967 from CheXpert and 39,875 from PadChest. Since the annotations of the findings on the source databases presented differences, some decisions were made to properly combined them. Specifically, ChestX-ray14 and CheXpert contained radiographs with 14 findings, but not all of them were coincidental. For uniformity, infiltration and edema were merged, and pneumonia was merged with consolidation. Support devices class was suppressed. The final list included 16 findings, which were atelectasis, cardiomegaly, consolidation, edema, emphysema, enlarged cardiomediastinum, fibrosis, fracture, hernia, lung opacity, mass, nodule, pleural effusion, pleural affectation, pleural thickening and pneumothorax. For the PadChest dataset, a search of the images with the 16 resulting findings was performed and combined. Additionally, radiographs in Lateral view were excluded.

The different number of samples per finding is shown in Table 1. Since each radiograph could present more than one finding the sum of the numbers provided on Table 1 is significantly higher. The normal class was used to train the normal vs abnormal classification model.

**Table 1.**
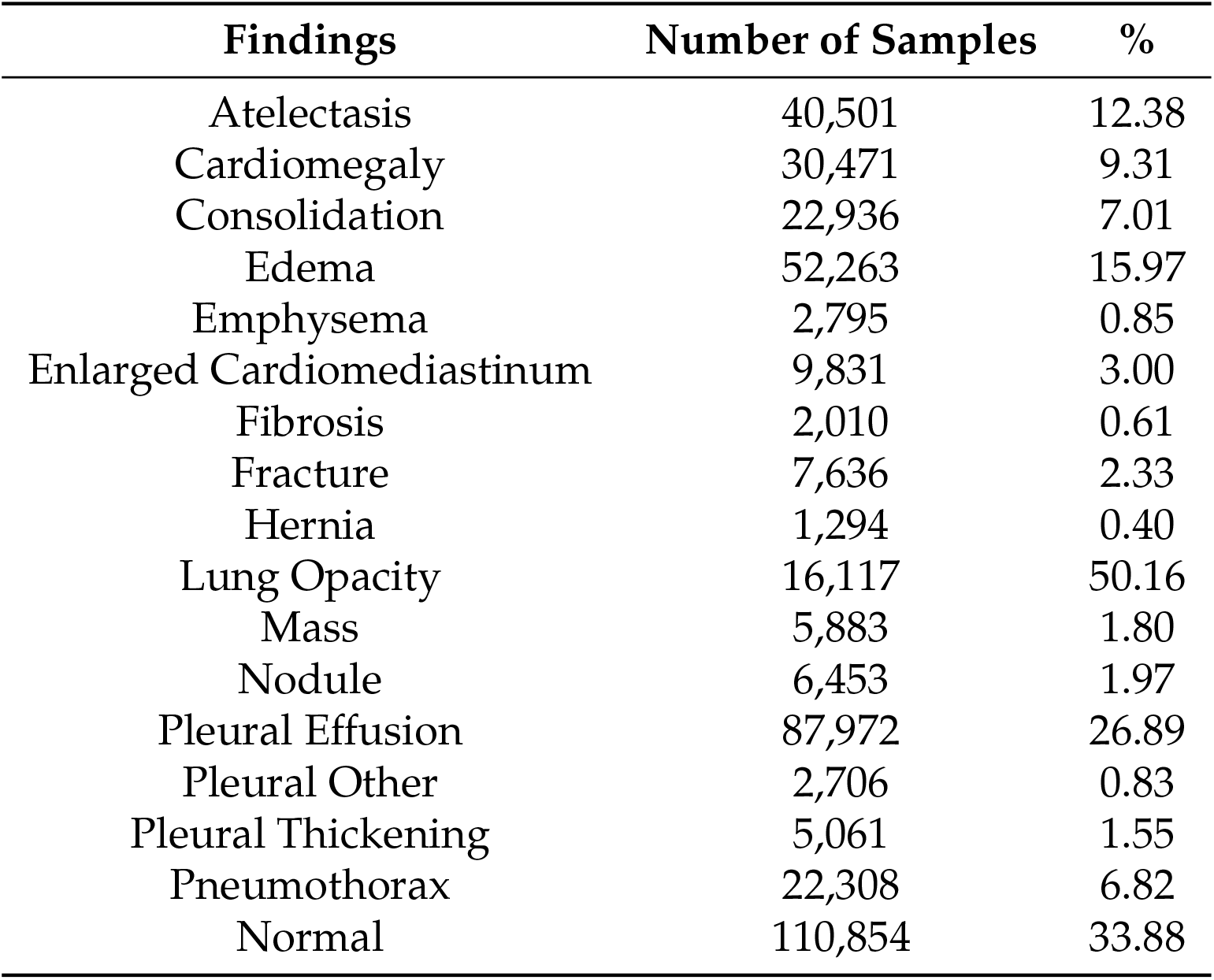
Distribution of the different findings

The database was divided in two partitions, the training set and the test set. To maximize the data for training the models, 323,776 (99%) were used for training and evaluation of the training process (validation) and 3,400 (1%) for testing the performance of the models. The training database was split in 16 different partitions designed to train finding-specific binary CNNs. Each partition was designed to contain all the samples of the dataset labeled with a specific finding as well as an equal number of samples without it randomly selected from the rest of classes. This split methodology guaranteed that balanced partitions were used to train the CNNs, as balanced datasets offer better performance than training a single multi-class CNN with a larger but imbalanced dataset (Buda et al., 2017). Another partition was created to train a referee fully connected neural network. Half of the images in this partition did not contain any abnormality and were labeled as normal, being all the other samples randomly selected from the radiographs with at least one of the 16 findings. Finally, the 17 partitions were split in training (80%) and validation (20%). Analogously, the test set was composed of 17 balanced partitions of 200 randomly selected radiographs as well.

The radiographs were prepared to improve the performance of the algorithm. The pipeline consisted of an initial histogram equalization, applied to all images to improve the contrast; then, intensity values were normalized to the range 0-1; and, finally, images were standardized using the mean and standard deviation of the training partitions to improve the convergence of the networks during training. The pixel intensity values of the radiographs were normalized using ImageNet statistics to improve compatibility with the pre-trained weights of the CNNs.

### 2.2. System’s pipeline

The development of the classifier algorithm included several concatenated steps (Figure 1). Initially, the three databases were combined, processed, and divided to train 16-binary pathology-specific CNN classifications. Once the 16 CNNs were trained, a novel approach for creating a triage system was implemented. In this step, CNNs’ outcomes were combined using a referee fully connected neural network that learnt to combine the predictions of the finding-specific CNNs to obtain the final abnormal probability, herein defined as ‘referee’ network. An evaluation of the trained models was carried out using the test set partition of the combined database, which was not used for training and contained 3,400 samples.

**Figure 1.**
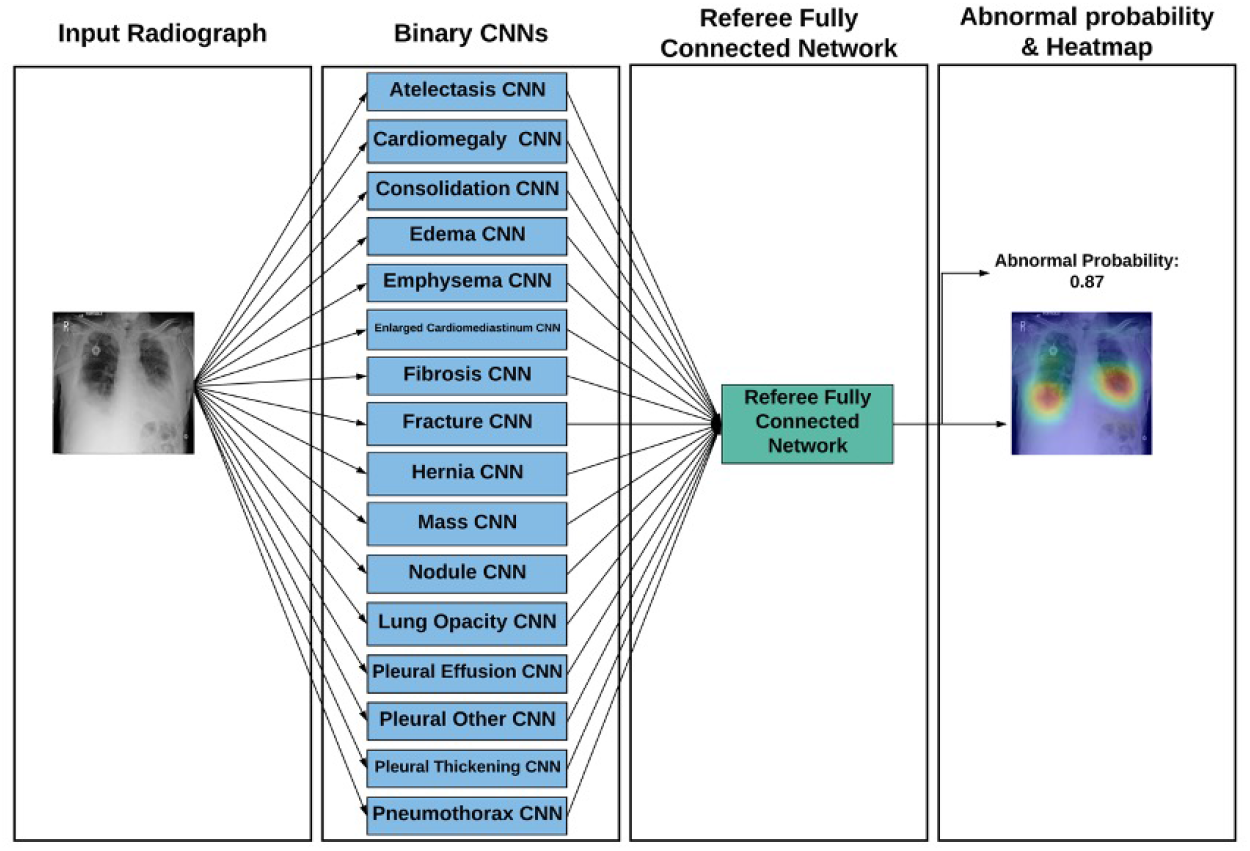
A visual diagram of the system’s pipeline

### 2.3. Training binary classifiers

The VGG-19 CNN architecture was used to train the 16 finding-specific models (Simonyan & Zisserman, 2014) (Figure 2), pre-trained with ImageNet. The output layer was modified to adapt the CNNs for the binary classification. The architecture was selected due to the higher dimensionality of its output feature maps if compared with ResNet (He et al., 2015) or DenseNet (Huang et al., 2017).

**Figure 2.**
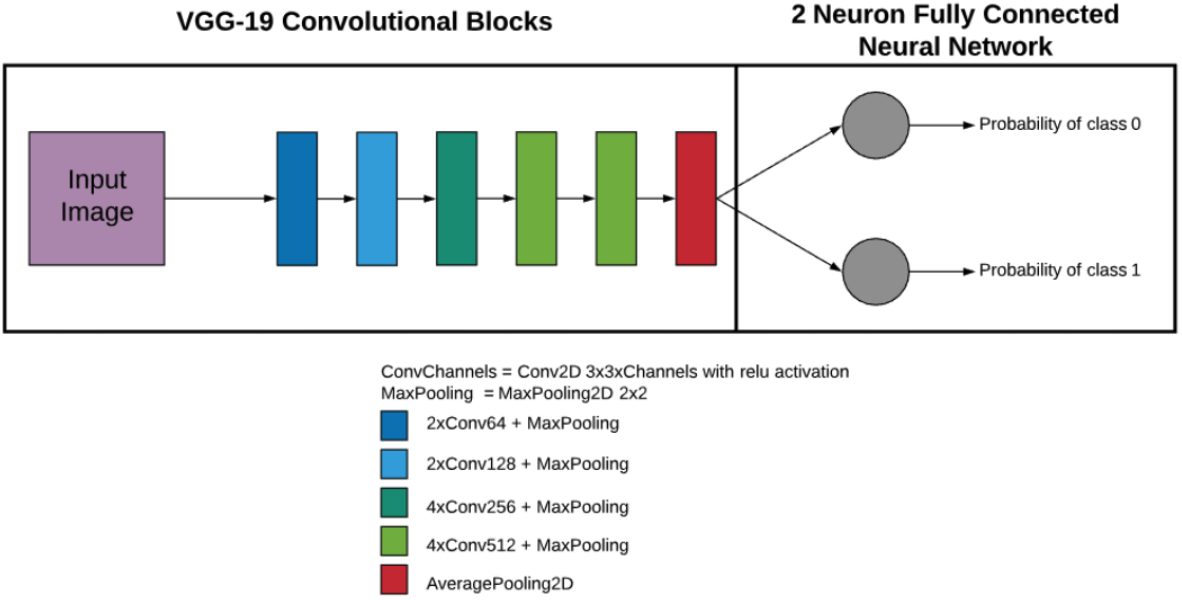
Architecture of the proposed CNN for finding-specific binary classification

The training process of the 16 CNN models consisted of 8 stages. During the first 7 stages, which lasted 5 epochs each, the convolutional layers were frozen in the odd stages and unfrozen in the even ones. The loss function used was categorical cross-entropy (CCE). The learning rate was customized for each stage and each CNN model independently ranging from 1.00E-08 to 1.00E-03. The image input size was increased along the stages starting at 3×128×128 until 3×320×320. During the training process data augmentation techniques were applied, which consisted of random rotations in the range of [-20, 20] degrees, zooms in the range [0.95, 1.05] and lighting variations in the range [0%, 10%].

The 16 finding-specific CNNs were used as feature extractors, providing the probability of each finding to be present on the radiograph. Thereafter, images of the normal vs abnormal partitions were processed by the CNNs to obtain their per-finding probabilities. These probabilities were then used as an input to the referee network, which was composed of 3 dense layers of 8, 4 and 2 neurons. The activation functions chosen were ReLU for the input and hidden layers and SoftMax for the output layer. The model was trained during 200 epochs using a learning rate of 1.00E-03 and CCE as loss function.

The metrics used to evaluate the statistics of the models’ performance were obtained by comparing the predictions of the models and the ground truth on the test set. These metrics were accuracy, precision, sensitivity, specificity, F1 score, and Area Under the Curve (AUC).

The models were trained using an NVIDIA QUADRO GP100 graphic card and an Intel Xeon SkyLake 6132. The deep learning framework used to train the models was Pytorch, and additionally the fastai library was used to train de finding specific CNNs.

### 2.4. Class activation maps

Class activation maps (Zhou et al., 2015) were used to provide information about the areas that were more influential in the outputs of the CNNs. A class activation map gets the discriminative image regions used by a CNN to identify a specific abnormality in the image. Adding a global average pooling between the convolutional and fully connected layers of the CNNs was needed to obtain these maps. The global average pooling layer allowed to directly connect each output feature map (f_k_) with an output class neuron (c) through a network weight (w_c,k_). In order to be able to calculate each class activation map (M_c_) a summatory of the different feature maps weighted by their respective class weight was performed by means of the following expression:

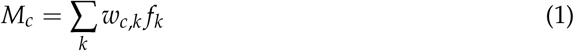

Due to the maxpooling layers of the proposed architecture (Simonyan & Zisserman, 2014), the output feature maps were smaller than the input image, therefore the class activation maps were resized to fit it. Several results of this technique can be seen in Figure 3.

**Figure 3.**
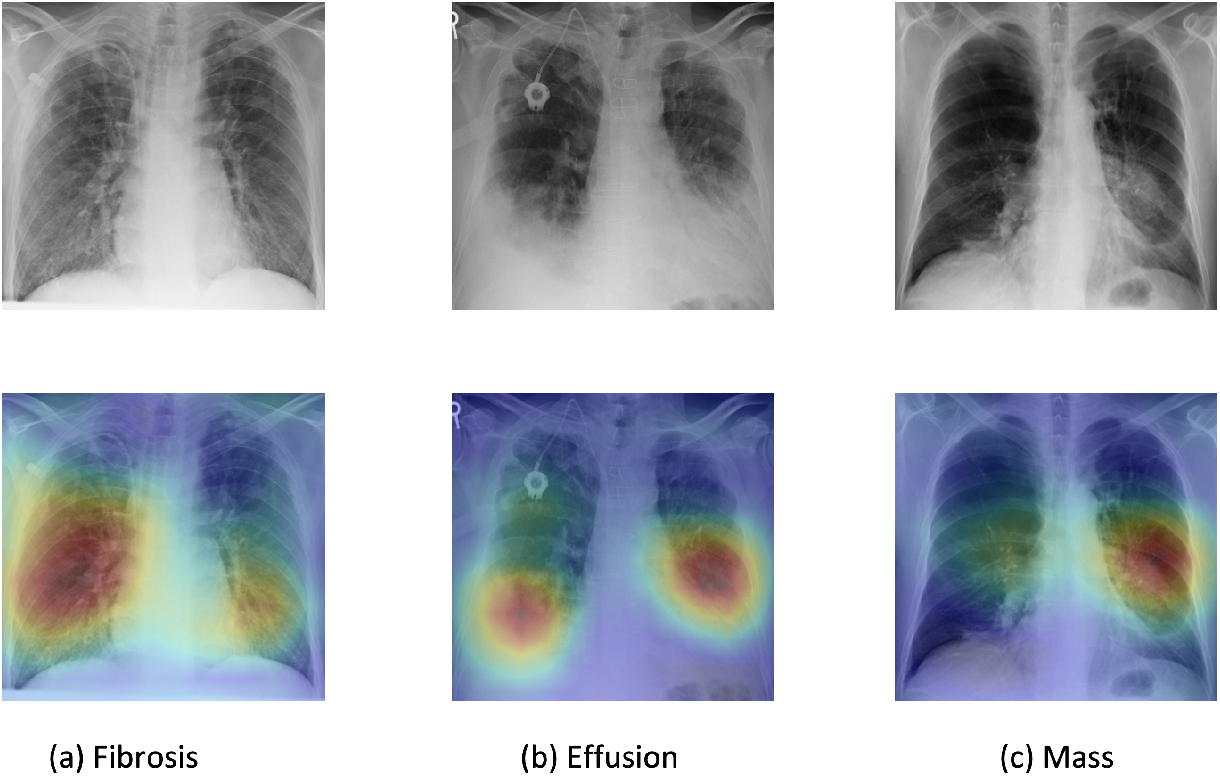
Class activation maps correctly overlapped with three different abnormalities. The upper row shows the input images to the finding-specific classifiers, the lower row represents the output class activation maps for the detected findings. Where: (a) presents reticular shadowing (fibrosis); (b) presents accumulation of fluid (pleural effusion); and (c) presents a large pulmonary opacity (Mass).

## 3. Results

The different metrics obtained using the 3,400 radiographs test set showed that the classifiers for the 16 different findings obtained different performances, due to the uneven size of the training datasets and to the different level of complexity of the visual patterns that represent the analyzed pathologies. The model trained for hernia classification obtained the best results with an AUC of 0.99. The different metrics obtained are shown in Table 2.

**Table 2.** Statistical information (accuracy, precision, sensitivity, specificity, F1 score, AUC) of the trained models.

| Findings | Accuracy | Precision | Sensitivity | Specificity | F1 Score | AUC | Threshold |
| --- | --- | --- | --- | --- | --- | --- | --- |
| Atelectasis | 0.82 | 0.79 | 0.87 | 0.77 | 0.83 | 0.88 | 0.40 |
| Cardiomegaly | 0.87 | 0.85 | 0.89 | 0.85 | 0.87 | 0.96 | 0.28 |
| Consolidation | 0.81 | 0.77 | 0.88 | 0.74 | 0.82 | 0.85 | 0.42 |
| Edema | 0.87 | 0.81 | 0.96 | 0.78 | 0.88 | 0.93 | 0.26 |
| Emphysema | 0.92 | 0.90 | 0.95 | 0.90 | 0.93 | 0.98 | 0.48 |
| Enlarged Cardiome-diastinum | 0.76 | 0.73 | 0.84 | 0.69 | 0.78 | 0.83 | 0.54 |
| Fibrosis | 0.86 | 0.80 | 0.97 | 0.76 | 0.88 | 0.94 | 0.34 |
| Fracture | 0.82 | 0.78 | 0.88 | 0.76 | 0.83 | 0.89 | 0.48 |
| Hernia | 0.92 | 0.88 | 0.97 | 0.87 | 0.92 | 0.99 | 0.36 |
| Lung Opacity | 0.84 | 0.87 | 0.81 | 0.88 | 0.84 | 0.91 | 0.62 |
| Mass | 0.84 | 0.79 | 0.93 | 0.75 | 0.85 | 0.91 | 0.48 |
| Nodule | 0.84 | 0.85 | 0.83 | 0.85 | 0.84 | 0.91 | 0.20 |
| Pleural Effusion | 0.83 | 0.82 | 0.85 | 0.81 | 0.83 | 0.89 | 0.52 |
| Pleural Other | 0.85 | 0.84 | 0.86 | 0.84 | 0.85 | 0.91 | 0.46 |
| Pleural Thickening | 0.87 | 0.85 | 0.89 | 0.85 | 0.87 | 0.94 | 0.56 |
| Pneumothorax | 0.80 | 0.79 | 0.82 | 0.79 | 0.81 | 0.90 | 0.51 |
| Normal | 0.77 | 0.70 | 0.97 | 0.58 | 0.81 | 0.94 | 0.20 |

**Table 3.**
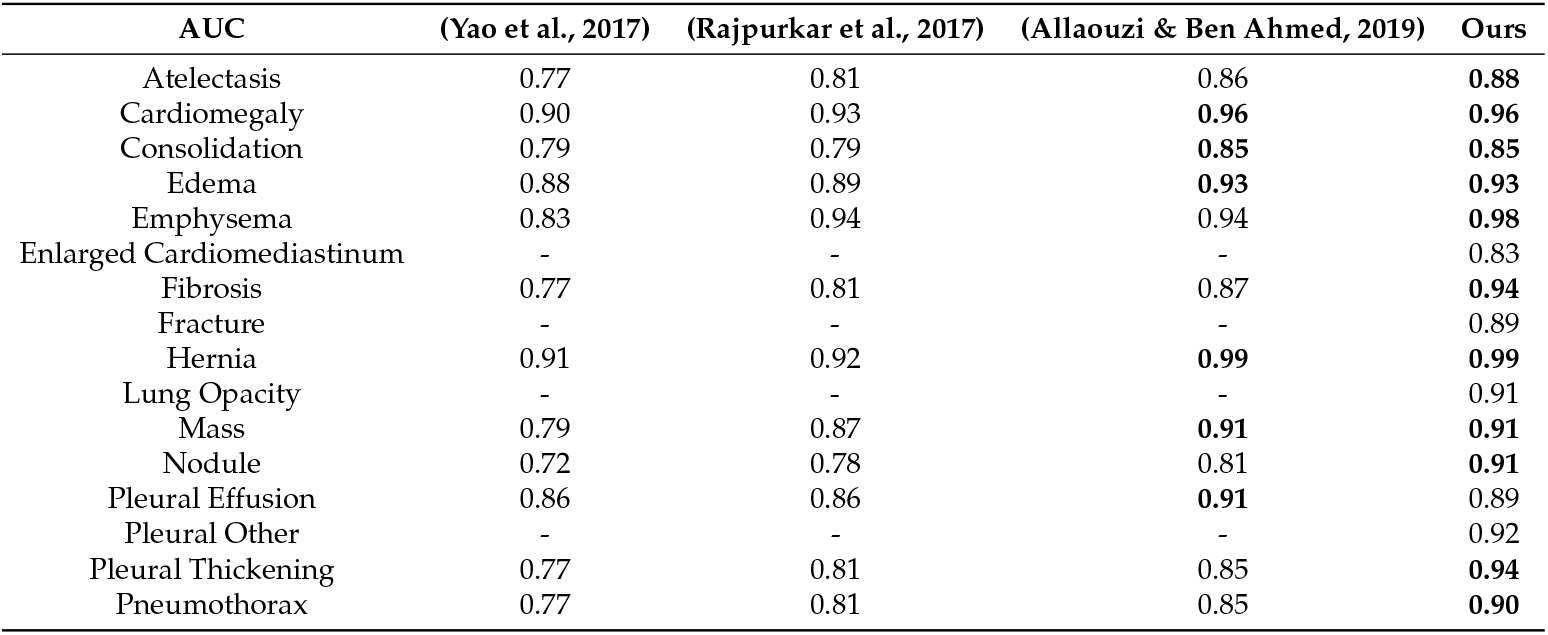
Comparison of the results obtained with our method and some prior works.

The process followed to determine the thresholds that are shown in the Table 2 for the 16 findings was the analysis of the accuracy, sensitivity and specificity obtained using all thresholds in between 0 and 1 with a step of 0.01. The criteria to select the threshold for all finding classifiers (except for normal vs abnormal classification) was to choose the point closest to the maximum accuracy value that presented a sensitivity higher than 0.80. The criteria to select the normal vs abnormal threshold was to choose a point with a sensitivity higher than 0.95 to reduce false negatives.

## 4. Discussion

This research proposes a novel approach for the automatic classification of chest radiographs based on a combination of CNNs by means of a referee neural network. The developed solution detects abnormal radiographs. This classification aims to accelerate the task of reporting plain radiographs, avoiding delays in diagnosis and improving patients’ outcomes. This prioritization of reports based on the probability of abnormal findings will help radiologists focusing on those radiographs more likely of being abnormal, helping to facilitate the radiologists’ recognition of abnormalities and avoiding medico-legal problems related to false results. Therefore, this tool will help to decrease the time to report abnormal radiographs. Although not proven, it is important to highlight that this classifier will adjust the radiologists’ efforts towards a more efficient environment. The advantages of using a chest X-ray classifier in clinical practice were previously demonstrated by implementing an AI system to predict the clinical priority of radiographs, which reduced the average reporting delay from 11 to 3 days for critical imaging findings and from 8 to 4 days for urgent imaging findings. The AI system utilized in the mentioned study was trained with a database of 470,388 adult chest radiographs which is not public as of now, therefore direct comparisons with this work are not possible (Annarumma et al., 2019).

Recent advancements in large public databases and DL have enabled the development of algorithms to help radiologists to evaluate plain X-rays. The publicly available imaging datasets ChestX-ray14 (Wang et al., 2017), CheXpert (Irvin et al., 2017) and PadChest (Bustos et al., 2020), which include a large number of chest radiographs labeled using NLP radiology reports, allowed to carry out this study.

The outcomes from this research are compared with three recent studies (Table 4), even though direct comparisons are not possible because the proposed work is trained and tested with a combination of three datasets and the results of the studies evaluated are obtained using ChestX-ray14 exclusively.

(Yao et al., 2017) developed a statistical two-stage neural network model that joins a compactly connected image encoder with a recurrent neural network decoder in order to make more accurate predictions. However, our chest X-ray classifier based on CNNs had better outcomes, outperforming (Yao et al., 2017) on all the diagnosis.

(Rajpurkar, 2017) published and developed an algorithm, called CheXNet, to detect pneumonia from radiographs. The proposed methodology was based in a very deep learning CNN (121 layers) for multi-class classification, previously trained with the ImageNet dataset and fine-tuned with ChestX-ray14 dataset afterwards. The authors concluded that CheXNet surpassed radiologist performance in detecting pneumonia when evaluating radiographs. This conclusion proves that technology could have similar performance as humans. In comparison, our algorithm performs better than CheXNet in the classification of all the evaluated findings. To be properly compared, both algorithms have to be validated with the same external unseen database. The task for which CheXNet is used is not oriented to detect alterations and assist radiologists on prioritizing abnormal cases but to depict pneumonia, not covering other usual clinical conditions in daily practice. The classifier presented in this study prioritizes the abnormal radiographs to be reported by radiologists, maximizing efficiency, helping in diagnosis and improving patient outcomes. (Allaouzi & Ben Ahmed, 2019) published a comparison of different methodologies to tackle the complexity of the multi-label nature of chest X-ray classification since label imbalance and inter-dependence increase significantly the complexity of this tasks. In this study binary relevance, label powerset and classifier chain approaches were evaluated on ChestX-ray14 and CheXpert databases, achieving binary relevance the best average AUC among the different findings. Our method performs equally or better on 10 of 14 findings that could be compared.

Focusing on normal vs abnormal classification, (Hwang et al., 2019) trained a CNN using a total of 87,696 cases (53,621 normal and 34,074 abnormal). In a validation with an external database of a 1,015 (486 normal and 529 abnormal) the system presented an AUC of 0.98 for normal vs abnormal classification which is superior to the AUC of 0.94 obtained by the proposed system, which may be due to their system just focuses on four types of image findings to consider an image abnormal: pulmonary malignant neoplasms, active tuberculosis, pneumonia and pneumothorax. On the other hand, our system takes into consideration 16 different findings which increases the variability of the task to be solved.

This study had some limitations that must be considered. First, radiographs’ labels of the three datasets were obtained using NLP techniques from radiological reports. Future studies could be based on prospectively annotated chest X-ray with the use of structured reporting techniques implemented in clinical routine. The reported performance of this methodology in the case of ChesX-ray14 was an overall F1 score of 0.90, therefore this caused that not all radiographs were correctly labelled (Wang et al., 2017). In DL the quality of the dataset is fundamental to obtain robust models, labelling errors could worsen the results offered by the trained models. Also, images were resized to 320×320 pixels to be processed by the CNNs in order to create faster classifiers that do not need high computing power hardware. Resizing the images might have an impact on the interpretation of the image as resolution and information is adjusted, especially in pathologies that present subtle changes in the radiographs such as small nodules and edema. Even more, this research was trained with posterior-anterior chest X-rays, what can be considered a limited approach as lateral views might be also obtained when examining thoracic structures to provide an improved report, even though these views do not increase the diagnosis accuracy for some pathologies (Kluthke et al., 2016). Finally, due to the large class imbalance of the dataset, the 16-binary classification CNNs have been trained with different numbers of dataset samples. As a consequence, some classifiers are more robust than others. This is the most important challenge to undertake to further refine the performance of the system, despite it is hard to solve due to the varied occurrence of the different pathologies.

Future work could be focused at improving these results by training CNNs with higher or even native spatial resolutions of input radiographs, better quality and larger datasets, and with more sources of information like lateral radiographs and clinical history. In summary, a classifier of normal vs abnormal chest radiographs using a CNN based methodology that extracts patterns related with different radiological findings has been developed to solve a clinical challenge. The study validates the feasibility of pretrained and fine-tuned deep CNN as an automated imaging feature extractor to train classifiers for radiology diagnosis. The positive results demonstrate both the viability and effectiveness of the novel approach of using a referee neural network to ensemble a group of binary CNNs trained with balanced datasets.

## Data Availability

The data used in the present can be publicly accessed following the references included.

## Funding

This research received no external funding.

## Institutional Review Board Statement

Not applicable.

## Informed Consent Statement

Not applicable.

## Conflicts of Interest

The authors declare no conflict of interest.

## Citation

López-González, R.; Sánchez-García, J.;Fos-Guarinos, B.; García-Castro, F. Alberich-Bayarri, A.; Soria-Olivas, E.; Muñoz-Núñez, C. and Martí-Bonmatí, L. Automated Chest Radiographs Triage Reading by a Deep Learning Referee Network. *Preprints* **2021**, 1, 0. https://doi.org/

## Publisher’s Note

MDPI stays neutral with regard to jurisdictional claims in published maps and institutional affiliations.

## Abbreviations

The following abbreviations are used in this manuscript:

CNN: Convolutional Neural Network
AUC: Area Under the Curve
CAD: Computer-Aided Diagnosis
DL: Deep Learning
GPU: Graphical Processing Unit
SGD: Stochastic Gradient Descent
CCE: Categorical Cross-Entropy
NIH: National Institute of Health

## Notes

### Competing Interest Statement

The authors have declared no competing interest.

### Funding Statement

There was no funding for the present work.

### Author Declarations

The databases used for the present study were ChestXray14, CheXpert and PAdChest. We got approval for using the three databases for research purposes after registration in the websites where the different databases are available. The ChestXray14 database was approved by the NIH Clinical Center ethical committee. The CheXpert database was approved by the Stanford Hospital ethical committee. The PadChest database was approved by the Hospital Universitario de San Juan ethical committee.

